# Correlates of the Post-Stroke cognitive impairment among patients with first-ever stroke admitted at tertiary hospitals in Dodoma, Tanzania: a prospective longitudinal study

**DOI:** 10.1101/2023.06.20.23291668

**Authors:** Baraka Alphonce, John Meda, Azan Nyundo

## Abstract

**Introduction:** Stroke patients develop cognitive impairment that, significantly impacting their quality of life, their families, and the community as a whole, but they are not given attention. This study aims to determine the prevalence and predictors of post-stroke cognitive impairment (PSCI) among adult stroke patients admitted to a tertiary hospital in Dodoma, Tanzania.

**Methodology:** A prospective longitudinal study was conducted at tertiary hospitals in the Dodoma region, central Tanzania. A sample size of 158 participants with the first stroke confirmed by CT/MRI brain aged ≥ 18 years met the criteria. At baseline, social-demographic, cardiovascular risks and stroke characteristics were acquired and then at 30 days, participants were evaluated for depression and apathy.. Descriptive statistics were summarised as continuous data reported as Mean (SD) or Median (IQR), and categorical data were summarised using proportions and frequencies. Univariate and multivariable logistic regression analysis were computed to determine predictors of PSCI

**Results:** Of 158 participants, the mean age was 58.7 years, 57.6% were female, and 80.4% of participants met the criteria for post-stroke cognitive impairment. After multivariable logistic regression, left hemisphere stroke (AOR: 5.798, CI: 1.030 – 32.623, *p* = 0.046), a unit cm^3^ increase in infarct volume (AOR: 1.064, 95% CI: 1.018 – 1.113, *p* = 0.007), and apathy symptoms (AOR: 12.259, CI: 1.112 – 89.173, *p* = 0.041) had a significant association with PSCI.

**Conclusion:** The study showed a high prevalence of PSCI; profiling at-risk stroke survivors in a timely intervention may improve their prognosis. Future studies in the area would inform future interventions and policies.

## Introduction

Stroke is the leading cause of death and disability, with about sixty-seven million people worldwide suffering their first stroke yearly, of which approximately 5,700,000 die and, 5000,000 are rendered incapacitated [1,2]. Stroke survivors develop cognitive impairment, which significantly impacts the quality of life of the sufferer, the family, and the community as a whole. PSCI is associated with reduced quality of life, increased likelihood of depressive symptoms, high dependence [3], increased health care cost, lost wages, and social isolation [4–6].

Globally, PSCI prevalence ranges from 35 to 92% [7–9]. In the few studies conducted in Sub-Saharan Africa, 40% prevalence between 3-months and 1-year and 34% prevalence at two years were observed in Nigerian and Ghanaian stroke survivors, respectively,[10,11]. The differences in the diagnostic tools used to assess PSCI in different studies, the timing of screening for cognitive impairment following a stroke, ethnicity, and cultural background could all explain the discrepancy in the prevalence of PSCI across settings [12].

Key variables predict PSCI at different stages of stroke; these include, Previous research has linked increasing advanced age, female gender, fewer years of formal education, hypertension, diabetes, dyslipidemia, atrial fibrillation, current alcohol consumption and cigarette smoking, stroke severity on admission, type of stroke, structures involved by stroke, stroke laterality, the volume of infarct or hematoma and neuropsychiatric manifestations at baseline[13–17]. The study aimed to assess the prevalence and predictors of PSCI in the early phase following a first stroke episode among patients admitted at tertiary hospitals in Dodoma, Tanzania.

## Material and Methods

### Study design and setting

This prospective longitudinal study design was conducted in Dodoma Referral Regional Hospital and Benjamin Mkapa Hospital, serving 20 – 30 stroke patients per month. Both are the designated teaching hospitals for the University of Dodoma for medical training at both undergraduate and residency level. With its well-built and state-of-the-art infrastructure, the Benjamin Mkapa Hospital is equipped with neuroimaging services, such as CT scans and MRI, serving Dodoma region, which is a capital city of Tanzania.

### Sample size and sampling procedure

Using a formula for proportion in a prospective cohort study [18] a minimum sample size of 143 was calculated. A total of 158 samples were collected over fifteen months, from June 2021 to September 2022. Participants who were readily accessible, willing to participate, and met the inclusion criteria were recruited until the required sample size was attained.

### Inclusion criteria/exclusion criteria

Patients included in the study were aged 18 years and above, who gave informed consent or proxy consent from a close relative in case the patient is incapable, presented with the first stroke within 14 days, and confirmed by CT scan or MRI brain. Those with previous history of stroke evidenced by clinical history or brain CT/MRI scans, traumatic intracerebral haemorrhage, intracerebral haemorrhage tumour, severe sensory impairment (blindness and deafness), TIA, subarachnoid haemorrhage and previous neurological disorder such as epilepsy, and patients with severe motor impairment in their dominant side were excluded.

### Outcome variable

Post-stroke cognitive impairment was defined as having a score of <23/30 on the MoCA, this score has better diagnostic accuracy than the commonly used 26/30 cut-off [19] and is useful in a less educated population. Translated and used in Swahili and. the tool examines eight major cognitive domains: visuospatial-executive (trail making B task, 3-D cube copy and clock drawing); naming (unfamiliar animals); language (sentence repetition and phonemic fluency task); short-term memory (delayed recall of words); abstraction (verbal abstraction); attention and calculation (digits forward and backwards, target detection using tapping, serial 7s subtraction) and orientation (time place and people) [20].

### Independent variables

Through a questionnaire that was structured based on evidence, variables such as age, gender, level of education, history of current alcohol use, history of current cigarette smoking, diabetes mellitus (defined as a history of confirmed diabetes melltus or the use of diabetic medications) [21] were acquired. Blood pressure (BP) readings were recorded according to the 2018 AHA/ACC Hypertension guideline for standard measurement of BP [22]. Hypertension was defined as BP ≥140/90 mmHg, or a patient on antihypertensive medications [23], radial pulse and heart rate was measured; a deficit of ten or more was considered indicative of atrial fibrillation [24].

A blood sample was analysed for Lipid profiles; according to the National Cholesterol Education Program (NCEP), dyslipidemia will be defined as HDL-Cholesterol <40 mg/dl and TC, LDL-Cholesterol and TG levels ≥200, ≥130 and ≥130 mg/dl, respectively [25]. Hyperglycemia was defined according to American Diabetes Association [26] for non-diabetic patient’s hyperglycemia was defined as random blood sugar >11.1 mmol/L, or fasting blood sugar > 7.0 mmol/L and diagnosis of diabetes was confirmed with a fasting blood sugar ≥ 7.0 mmol/L, or random blood glucose ≥ 11.1 mmo/L plus symptoms of hyperglycaemia or glycated haemoglobin≥ 6.5 % A 12-lead ECG was done on each participant under the supervision of a consultant cardiologist. Atrial Fibrillation was diagnosed by the absence of P waves and irregular-irregular RR interval [27]. A 24-hour ECG Holter was done in a patient with ischemic stroke whose 12-lead ECG tracing was normal to screen for paroxysmal atrial fibrillation [28].

All patients had a CT scan (SIEMENS-SOMATOM Definition Flash) or MRI brain model MAGNETUM SPECTRA A TIM +Dot System 3T, sequences 3D-T1, axial T2, 3D-FLAIR, DWI. Strokes were characterised according to type, hemisphere affected, cortical or subcortical, and the ellipsoid method measured the volume of infarct/hematoma. [29,30]. All patients were evaluated for renal function status before undergoing brain imaging to reduce the risk of contrast-induced nephropathy [31].

The Patient Health Questionnaire (PHQ) – 9, with a total score of 27, was used to screen stroke survivors for post-stroke depression; the score was classified as (1 - 4) minimal depression, (5 – 9) mild depression, (10 – 14) moderate depression, (15 – 19) moderately severe depression, and (20 – 27) severe depression. A study done in Tanzania proved that the PHQ-9 is the right tool for screening depressive symptoms in primary healthcare settings. The results favour a cut-off score of greater than or equal to 9, with a sensitivity of 78% and a specificity of 87% for depressive symptoms equivalent to a major depressive episode [32]. Apathy was identified using the apathy evaluation scale, a score > 38 which has sensitivity and specificity around 80 % and 100%, respectively [33], was suggestive of apathy symptoms [34].

### Data analysis

For statistical analysis, data were entered on a Microsoft Excel sheet and then converted to IBM SPSS PC version 26. Continuous variables were reported as mean and standard deviation (SD), or Median and interquartile ranges; frequencies and percentages were used for categorical variables. Chi-square, and Mann-Whitney U test were used to determine the difference between independent variables by post-stroke cognitive outcomes; these included, Social-Demographic, cardiovascular risk factors, stroke characteristics, and neuropsychiatric manifestations, which are depression and apathy.To determine variables to use for adjustment, the predictors were evaluated by binary logistic regression and only variables that met a 20% (p-value≤0.2) statistical significance [35] were selected for multivariable Logistic regression analysis to determine independent predictors for post-stroke cognitive impairment. The adjusted odds ratio (aOR) and the 95 % confidence interval (CI) was determined. Statistical significance was determined by a two-sided p ≤ 0.05.

### Ethical issues

The Vice Chancellor’s office at the University of Dodoma permitted the study after obtaining ethical clearance from the Directorate of Research and Publications (reference number MA.84/261/02). The administrative departments of Benjamin Mkapa and Dodoma Regional Referral Hospitals approved data collection with reference numbers AB.150/293/01/196 and EB.21/267/01/123, respectively. Participants were informed that their participation was fully optional and that they might opt out at any moment and their standard of care was unaffected by their decision to participate. Participants’ names were substituted with identification numbers to ensure privacy and confidentiality. Stroke survivors who had depressed symptoms were referred to a psychiatrist for additional examination and treatment. Thereafter, participants were asked to provide a written or verbal informed consent under the witness of the close relative. For those who could not provide informed consent, a custodian who had to be a close family member or guardian provided the assent on behalf.

## Results

In this study, 255 stroke patients were evaluated for eligibility criteria (Fig.1). 158 participants with first-ever stroke were evaluated for the outcome of interest at 30 days of follow-up, among them, one hundred and twenty-seven (80.4%) had cognitive impairment.

**Figure 1:**
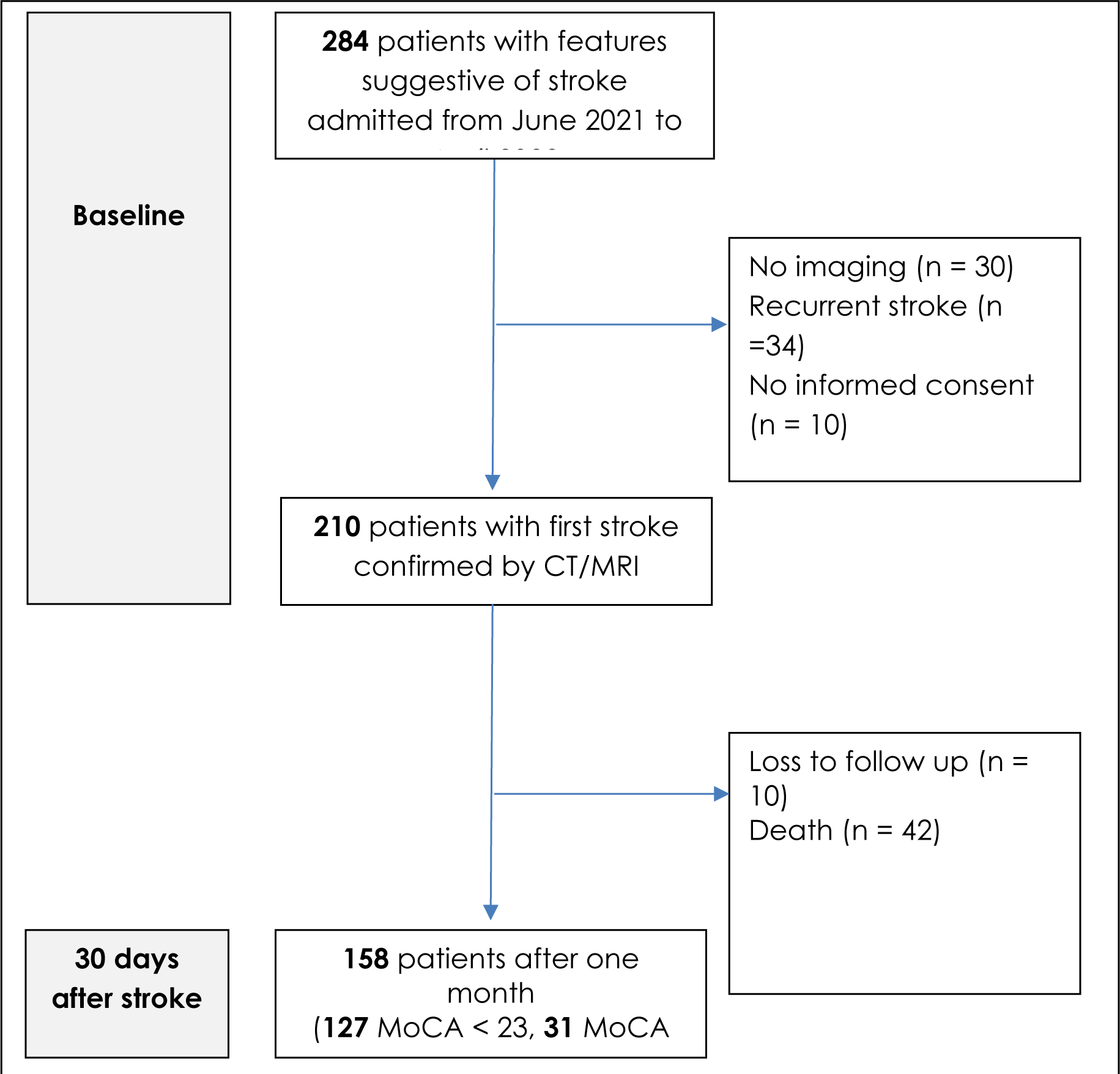
Algorithm for enrolment of study participants and 30 days post-stroke cognitive outcome.

**Fig 2.**
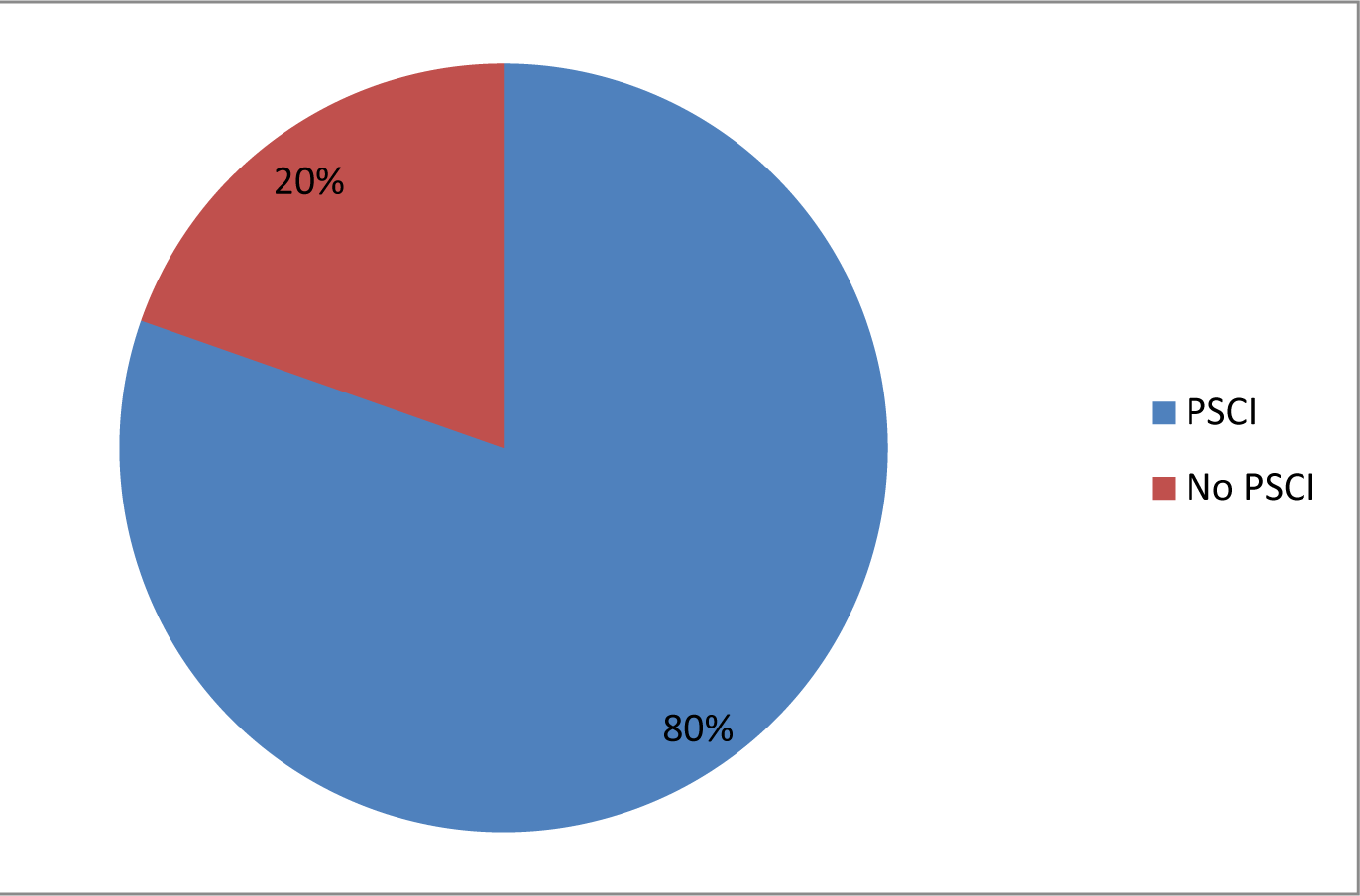
Pie chart demonstrating the prevalence of PSCI at one month, N(158)

### Social Demographic characteristics

Of the 158 study participants, (57.6%) were females, and the population mean age was 58.7 ± 13.4 years. Approximately (49.4%) attained seven or less years of formal education, 50%residing in urban regions, and the majority (66.5%) were referred from a primary health care facility. As for socio-demograohic factors, only older age (*p* > 0.001), and seven or fewer years of formal education (*p* < 0.001) showed significant differences with post-stroke cognitive outcomes (Table 1)

**Table 1.** Demographic, clinical characteristics and vascular risk factors of patients with different cognitive outcomes (N=158)

|  | All<br>(N=158) | No PSCI (N<br>= 31) | PSCI (N = 127) |  |
| --- | --- | --- | --- | --- |
| Variables | Frequency<br>(%) | Frequency<br>(%) | Frequency<br>(%) | Significance |
| **Social Demographic characteristics** |  |  |  |  |
| Age (**Mean ± SD**) | 58.7 ± 13.4 | 50.5 ± 12.5 | 60 ± 12.9 | |
| <50 | 37 (23.4) | 10 (32.3) | 27 (21.3) | 0.001 |
| 50 - 60 | 53 (33.5) | 17 (54.8) | 36 (28.3) |  |
| >60 | 68 (43.1) | 4 (12.9) | 64 (50.4) |  |
| Sex |  |  |  |  |
| Male | 67 (42.4) | 9 (29) | 58 (45.7) | 0.093 |
| Female | 91 (57.6) | 22 (71) | 69 (54.3) |  |
| Residence |  |  |  |  |
| Urban | 79 (50) | 11 (35.5) | 68 (53.5) | 0.071 |
| Rural | 79 (50) | 20 (64.5) | 59 (46.5) |  |
| Referral status |  |  |  |  |
| Self | 53 (33.5) | 13 (41.9) | 40 (31.5) | 0.270 |
| Referred | 105 (66.5) | 18 (58.1) | 87 (68.5) |  |
| Years of formal education |  |  |  |  |
| ≤ 7 years | 78 (49.4) | 4 (12.9) | 74 (58.3) | < 0.001 |
| ≥ 8 years | 80 (50.6) | 27 (87.1) | 53 (41.7) | |
| **Vascular risk factors** |  |  |  |  |
| Current Cigarette smoking | 33 (20.9) | 5 (16.1) | 28 (22) | 0.467 |
| Current Alcohol intake | 27 (17.1) | 1 (3.2) | 26 (20.5) | 0.022 |
| Hypertension | 117 (94.1) | 19 (61.3) | 98 (77.2) | 0.071 |
| Diabetes | 36 (22.8) | 7 (22.6) | 29 (22.8) | 0.976 |
| Atrial fibrillation | 31 (19.6) | 4 (12.9) | 27 (21.3) | 0.294 |
| Dyslipidaemia | 106 (67.1) | 20 (64.5) | 86 (67.7) | 0.734 |
| **Stroke characteristics** |  |  |  |  |
| NIHSS scale: **Median (IQR)** | 12 (7) | 12 (8) | 12(7) | 0.813 |
| Stroke type |  |  |  |  |
| Ischemic | 88 (69.3) | 21 (67.7) | 88 (69.3) | 0.867 |
| Haemorrhagic | 39 (30.7) | 10 (32.3) | 39 (30.7) |  |
| Infarct volume (cm<sup>3</sup>): **Median (IQR)** | 40 (87) | 9 (26) | 50 (84) | < 0.001 |
| Hematoma volume (cm<sup>3</sup>): **Median (IQR)** | 20.7 (28) | 15 (25) | 23 (35) | 0.248 |
| Structures involved |  |  |  |  |
| Cortical | 88 (55.7) | 10 (32.3) | 78 (61.4) | 0.003 |
| Subcortical | 70 (44.3) | 21 (67.7) | 49 (38.6) |  |
| Stroke laterality |  |  |  |  |
| Left | 97 (61.4) | 10 (32.3) | 87 (68.5) | < 0.001 |
| Right/brain stem, cerebellum | 61 (38.6) | 21 (67.7) | 40 (31.5) |  |
| Stroke vascular territory |  |  |  |  |
| Posterior | 16 (10.1) | 5 (16.1) | 11 (8.7) | 0.217 |
| Anterior | 142 (89.9) | 26 (83.9) | 116 (91.3) |  |
| **Emotional factors** |  |  |  |  |
| Apathy | 57 (36.1) | 3 (9.7) | 54 (42.5) | 0.001 |
| AES score: **Median (IQR)** | 34 (17) | 16 (13) | 36 (14) |  |
| Depression |  |  |  |  |
| PHQ-9: **Median (IQR)** | 8 (10) | 3 (4) | 9 (9) |  |
| Minimal-moderate | 127 (80.4) | 27 (87.1) | 100 (78.7) | 0.294 |
| Severe | 31 (19.6) | 4 (12.9) | 27 (21.3) |  |

### Clinical characteristics of participants

The majority of the participants, 117 (94.1 %), had hypertension, 36 (22.6%) were diabetic, 106 (67.1 %) had dyslipidaemia, 31 (19.6%) had atrial fibrillation while a minority, 33 (20.9%) were cigarette smokers and 27 (17.1%) had a history of alcohol use. A larger proportion (20.5%) of participants with a history of alcohol use were significantly overrepresented among stroke survivors with post-stroke cognitive impairment (*p* = 0.022); otherwise, there was no significant difference in post-stroke cognitive outcomes by other vascular risk factors (Table 1)

The median NIHSS score of all participants was 12, IQR (7.0). The majority (69.3%) had an ischemic stroke, while the median volume of infarct and hematoma was 40, IQR (87) and 20.7, IQR (28), respectively. In terms of stroke characteristics, 97 (61.5 %) had left hemisphere stroke, 142 (89.9%) had anterior circulation involvement, and 88(55.6%) involved primary cortical structures versus 70 (44.3), which had subcortical involvement. Among the stroke characteristics, only infarct volume (*p* < 0.001), cortical strokes (*p* = 0.003), and left-sided strokes (*p* < 0.001) had significantly higher proportions among participants with post-stroke cognitive impairment (Table 1).

Most of the participants (80.4%) met the criteria for minimal to moderate depression with the median score of 8, IQR (10 for PHQ-9 while apathy was found in (36.1%) of participants, with median EAS score 34, IQR (17). Only apathy was significantly overrepresented among participants with post-stroke cognitive impairment (*p* < 0.001) (Table 1).

### Predictors of post-stroke cognitive impairment

On unadjusted logistic regression, increasing age, less than eight years of formal education, hypertension, history of current alcohol use, increasing infarct volume, left-sided stroke, cortical stroke, and apathy were all significantly associated with post-stroke cognitive impairment (Table 2). However, under adjusted logistic regression, only a unit cm^3^ increasing infarct volume (AOR: 1.064, 95% CI: 1.018 – 1.113, *p* = 0.007), left-sided stroke (AOR: 5.798, CI: 1.030 – 32.623, *p* = 0.046), and apathy (AOR: 12.259, CI: 1.112 – 89.173, *p* = 0.041) remained significantly associated with cognitive impairment at p≤0.05 level of significance while increasing age (*p* = 0.072) was significant at 10% level of significance, (Table 2).

**Table 2:**
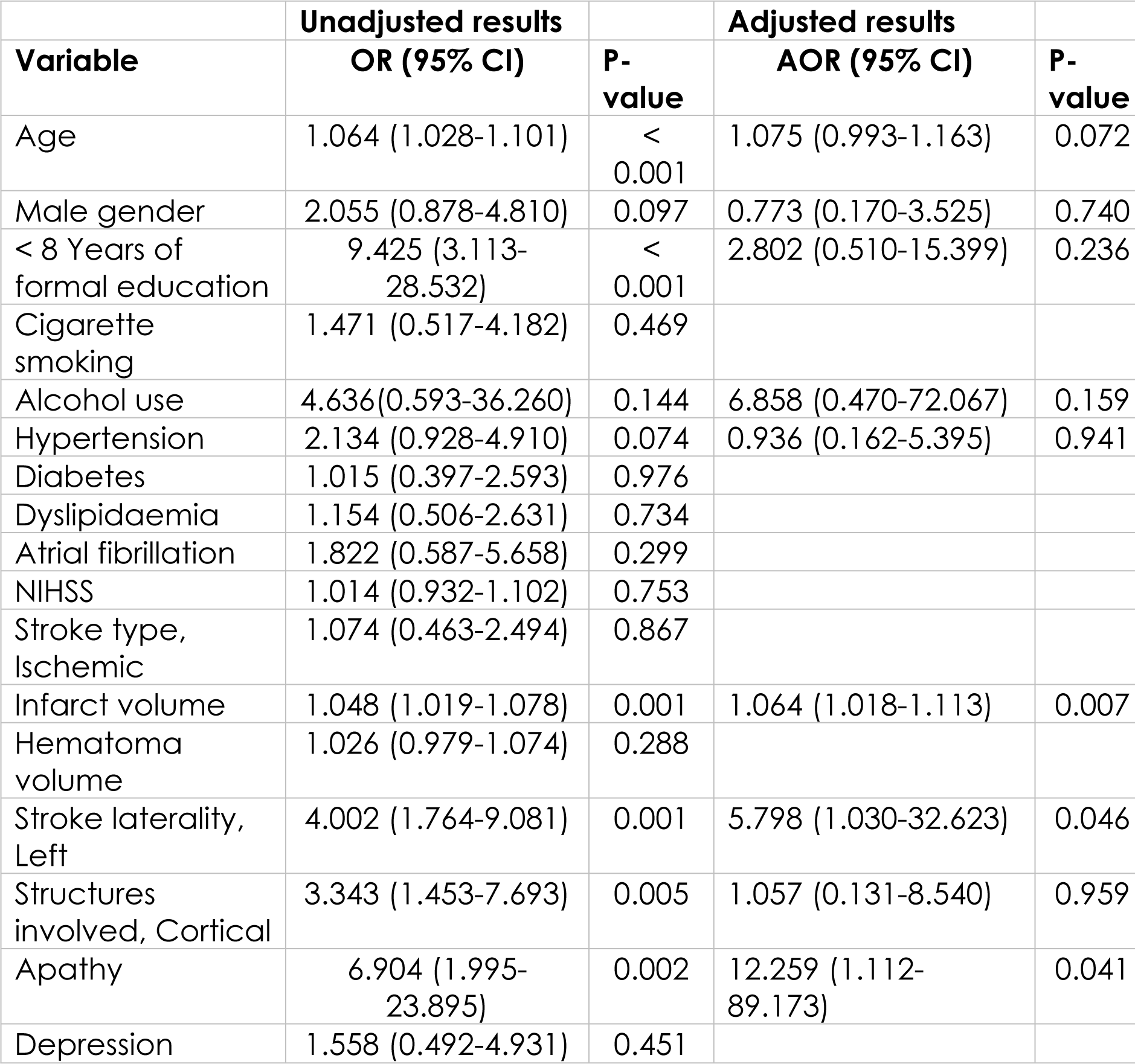
Logistic regression analysis of predictors of cognitive Impairment at 1-month.

## Discussion

The main objective of this study was to determine the predictors of early cognitive impairment among patients with the first episode of stroke admitted at tertiary hospitals in Dodoma. We revealed a high prevalence of PSCI at 30 days (80.4%), which was independently associated with stroke laterality, increasing infarct volume and apathy. Moreover, we also determined the prevalence of post-stroke cognitive impairment.

Depending on the definition, stages of stroke, stroke severity at admission, population heterogeneity, and regions, the prevalence of PSCI has been reported to range from 20-70% [7–9]. Studies screened with MoCA report a PSCI prevalence of 57-67% at an acute phase among individuals without pre-morbid cognitive impairment [36]. The prevalence of PSCI increases to 66.4% and 75.2% two to eight weeks after the stroke if the history of previous stroke is not excluded [7,9,37]. However, lower prevalence has been observed when comprehensive neuropsychological batter is used; for example, studies in Ghana and Nigeria showed 34% and 39% prevalence of PSCI, respectively [10,11]. While the PSCI prevalence varies globally, our study supports previous observations of high incidence and prevalence of PSCI in the early phase following an episode of stroke [38]. Although the study population’s mean age of 58.7 may anticipate a relatively lower prevalence compared to previous studies with an older population of 60 years and above, not excluding participants with prior history of cognitive impairment may raise the PSCI prevalence [7,9], furthermore the majority of the participants had lower lever of formal educations and significant proportion residing from rural settings, these two factors may predict poor performance of neurocognitive assessment [10].

Similar to previous studies that used the same assessment tool, left hemisphere stroke was a significant predictor of PSCI at one month [39] and even six months [40]. The potential of left hemisphere stroke to influence cognitive performance acutely reflects that language is a left hemispheric cognitive domain for more than 90% of the population worldwide [41]. Language is a key domain in the MoCA assessment [42] and the screening tool has a relatively lower sensitivity (72%) for identifying right-hemispheric cognitive deficits compared to 94% sensitivity for left-hemispheric cognitive deficits [43].

Our study reveals that a unit (cm^3^) increase in infarct size was a significant predictor of PSCI. The relationship between infarct volume and PSCI was first reported by Tomlison et al, [44]who found that infarct volume closer to 100 cm^3^ increased the risk of PSCI significantly while Kumral et al [45] also showed that infarct volume greater than 90 cm^3^ is an independent predictor of PSCI. Although methodological diversity in measuring and calculating infarct volume could explain from differences in findings across settings [46], our findings are generally consistent with observations that increased of an infarct volume of even greater than 17cm^3^ may be an independent predictor of PSCI.

In this index study, 36.1% of the subjects in our study met a criteria for apathy, while lower prevalence (28%) has been reported among patients with chronic stroke [47], a similar rate of 36% is reported in a metanalysis where symptoms of apathy are independent predictor of PSCI [14,48]. The higher prevalence of apathy in our study can be attributed to the higher sensitivity of the AES > 38 cut-off [48]and the earlier time of apathy assessment [49]. The same underlying brain injury may produce apathy and cognitive impairment; in particular, the frontal lobes and associated subcortical structures that are thought to be involved in apathy are also linked to various cognitive functions [50]; additionally, loss of cognitive capacities is likely to limit a person’s ability to organise goal-directed behaviour. According to this viewpoint, apathy is an inherent symptom (or indication) of cognitive impairment rather than a unique neuropsychiatric illness.

Early stratification of risk for cognitive impairment after stroke is strongly supported by the link between early cognitive dysfunction after stroke and overall functional status, as well as the level of morbidity linked to cognitive impairment after stroke in the follow-up phase [51–53].

Even though there is not much evidence yet, pharmacological treatments, memory-improving techniques, and kinematic analysis are all possible ways to help people with cognitive impairment. This is because it is challenging for people with cognitive impairment to adhere to prescriptions and a healthy lifestyle, thus affecting their overall health [54]. It is therefore, essential to identify patients who would benefit from early cognitive assessment, intervention, and counselling on the odds of improvement.

### Limitation of the study

This was a longitudinal cohort study; there was no comparable non-stroke group to compare cognitive outcomes to make robust inferences. Cognitive functioning was assessed only once during the stroke recovery period, and pre-stroke cognitive performance was unknown. Thus, a survey such as an Informant Questionnaire for Cognitive Decline in the Elderly (IQCODE) [55] be included in research designs to collect an indicator of pre-stroke cognitive function. The use of MoCA instead of the gold standard comprehensive neuropsychological battery limited the diagnosis of participants with PSCI for whom MoCA may limited to classify accurately [58], nonetheless, MoCA has demonstrated a good screening capacity where assessment with gold standard tool is practically not possible.

## Conclusion

Early cognitive impairment is common in stroke survivors with unknown pre-morbid cognitive performance; however, stroke laterality, increasing infarct volume, and apathy are strong predictors of poor cognitive performance and may allow us to better identify and target at-risk individuals for aggressive rehabilitation in the acute setting. Well-designed studies are needed in sub-Saharan regions to understand better the magnitude, determinants, and characteristics of PSCI.

## Data Availability

All relevant data are within the manuscript and its Supporting Information files.

## Data Availability

All relevant data are within the manuscript and its Supporting Information files.

## References

1. Kulesh A, Drobakha V, Kuklina E, Nekrasova I, Shestakov V. Cytokine Response, Tract-Specific Fractional Anisotropy, and Brain Morphometry in Post-Stroke Cognitive Impairment. Journal of Stroke and Cerebrovascular Diseases. 2018 Jul 1. 27(7):1752–9. doi: 10.1016/j.jstrokecerebrovasdis.2018.02.004

2. Zhu Z, Chen L, Guo D, Zhong C, Wang A, Bu X, et al. Serum Rheumatoid Factor Levels at Acute Phase of Ischemic Stroke are Associated with Poststroke Cognitive Impairment. Journal of Stroke and Cerebrovascular Diseases. 2019 Apr 1. 28(4):1133–40. doi: 10.1016/j.jstrokecerebrovasdis.2018.12.049

3. Jeffares I, Rohde D, Doyle F, Horgan F, Hickey A. The impact of stroke, cognitive function and post-stroke cognitive impairment (PSCI) on healthcare utilisation in Ireland: a cross-sectional nationally representative study. BMC Health Services Research. 2022. 22(1):1–13. doi: 10.1186/s12913-022-07837-2

4. Fride Y, Adamit T, Maeir A, Ben Assayag E, Bornstein NM, Korczyn AD, et al. What are the correlates of cognition and participation to return to work after first ever mild stroke. Topics in Stroke Rehabilitation. 2015. 22(5):317–25. doi: 10.1179/1074935714Z.0000000013

5. Lees RA, Hendry BA K, Broomfield N, Stott D, Larner AJ, Quinn TJ. Cognitive assessment in stroke: feasibility and test properties using differing approaches to scoring of incomplete items. International Journal of Geriatric Psychiatry. 2017. 32(10):1072–8. doi: 10.1002/gps.4568

6. Pollock A, St George B, Fenton M, Firkins L. Top ten research priorities relating to life after stroke [Internet]. Vol. 11, The Lancet Neurology. Lancet Neurol; 2012. p. 209. doi: 10.1016/S1474-4422(12)70029-7

7. Gong L, Gu Y, Yu Q, Wang H, Zhu X, Dong Q, et al. Prognostic Factors for Cognitive Recovery Beyond Early Poststroke Cognitive Impairment (PSCI): A Prospective Cohort Study of Spontaneous Intracerebral Hemorrhage. Frontiers in Neurology. 2020. 11(April):1–10. doi: 10.3389/fneur.2020.00278

8. Nys GMS, Van Zandvoort MJE, De Kort PLM, Jansen BPW, De Haan EHF, Kappelle LJ. Cognitive disorders in acute stroke: Prevalence and clinical determinants. Cerebrovascular Diseases. 2007 May. 23(5–6):408–16. doi: 10.1159/000101464

9. Sharma R, Mallick D, Llinas RH, Marsh EB. Early Post-stroke Cognition: In-hospital Predictors and the Association With Functional Outcome. Frontiers in Neurology. 2020 Dec 23. 11:1817. doi: 10.3389/fneur.2020.613607

10. Akinyemi RO, Allan L, Owolabi MO, Akinyemi JO, Ogbole G, Ajani A, et al. Profile and determinants of vascular cognitive impairment in African stroke survivors: The CogFAST Nigeria Study. Journal of the Neurological Sciences. 2014. 346(1–2):241–9. doi: 10.1016/j.jns.2014.08.042

11. Sarfo FS, Akassi J, Adamu S, Obese V, Ovbiagele B. Burden and Predictors of Poststroke Cognitive Impairment in a Sample of Ghanaian Stroke Survivors. Journal of Stroke and Cerebrovascular Diseases. 2017. 26(11):2553–62. doi: 10.1016/j.jstrokecerebrovasdis.2017.05.041

12. Chi NF, Hu HH, Chan L, Wang CY, Chao SP, Huang LK, et al. Impaired cerebral autoregulation is associated with poststroke cognitive impairment. Annals of Clinical and Translational Neurology. 2020. 7(7):1092–102. doi: 10.1002/acn3.51075

13. Pendlebury ST, Rothwell PM. Prevalence, incidence, and factors associated with pre-stroke and post-stroke dementia: a systematic review and meta-analysis. The Lancet Neurology. 2009. 8(11):1006–18. doi: 10.1016/S1474-4422(09)70236-4

14. Douven E, Köhler S, Schievink SHJ, van Oostenbrugge RJ, Staals J, Verhey FRJ, et al. Baseline Vascular Cognitive Impairment Predicts the Course of Apathetic Symptoms After Stroke: The CASPER Study. American Journal of Geriatric Psychiatry. 2018 Mar 1. 26(3):291–300. doi: 10.1016/j.jagp.2017.09.022

15. Swardfager W, MacIntosh BJ. Depression, Type 2 Diabetes, and Poststroke Cognitive Impairment. Neurorehabilitation and Neural Repair. 2017 Jan 1. 31(1):48–55. doi: 10.1177/1545968316656054

16. Al Fawal B, Ibrahim A, Abd Elhamed M. Post-stroke dementia: frequency, predictors, and health impact. Egyptian Journal of Neurology, Psychiatry and Neurosurgery. 2021 Dec 1. 57(1):1–8. doi: 10.1186/S41983-021-00270-Y/FIGURES/3

17. Chander RJ, Lim L, Handa S, Hiu S, Choong A, Lin X, et al. Atrial Fibrillation is Independently Associated with Cognitive Impairment after Ischemic Stroke. Journal of Alzheimer’s Disease. 2017. 60(3):867–75. doi: 10.3233/JAD-170313

18. Baraka A, Meda J, Nyundo A. Predictors of post-stroke cognitive impairment at three-month following first episode of stroke among patients attended at tertiary hospitals in Dodoma, central Tanzania: A protocol of a prospective longitudinal observational study metadata. PLOS ONE. 2023 Mar 1. 18(3):e0273200. doi: 10.1371/JOURNAL.PONE.0273200

19. Carson N, Leach L, Murphy KJ. A re-examination of Montreal Cognitive Assessment (MoCA) cut-off scores. International Journal of Geriatric Psychiatry. 2018 Feb 1. 33(2):379–88. doi: 10.1002/GPS.4756

20. Julayanont, P., Phillips, N., Chertkow, H., and Nasreddine ZS. The Montreal Cognitive Assessment (MoCA): Concept and Clinical Review. 2012. (10).

21. Chen A, Akinyemi RO, Hase Y, Firbank MJ, Ndung’u MN, Foster V, et al. Frontal White matter hyperintensities, clasmatodendrosis and gliovascular abnormalities in ageing and post-stroke dementia. Brain. 2016 Jan 1. 139(1):242–58. doi: 10.1093/brain/awv328

22. Whelton PK, Carey RM, Aronow WS, Casey DE, Collins KJ, Himmelfarb CD, et al. 2017 ACC/AHA/AAPA/ABC/ACPM/AGS/APhA/ASH/ ASPC/NMA/PCNA guideline for the prevention, detection, evaluation, and management of high blood pressure in adults: Executive summary: A report of the American college of cardiology/American Heart Association task. Vol. 71, Hypertension. 2018. 1269–1324 p. doi: 10.1161/HYP.0000000000000066

23. Maduagwu SM, Umeonwuka CI, Mohammad HH, Oyeyemi AY, Nelson EC, Jaiyeola OA, et al. Reference Arm for Blood Pressure Measurement in Stroke Survivors. Middle East Journal of Rehabilitation and Health. 2018 Jan 30. 5(1). doi: 10.5812/mejrh.62368

24. Rajkumar A, Bhattacharjee A, Selvaraj RJ. Diagnostic accuracy of apex-pulse deficit for detecting atrial fibrillation. International Journal of Advanced Medical and Health Research. 2019. 6(2):52. doi: 10.4103/IJAMR.IJAMR_48_19

25. Cleeman JI. Executive summary of the third report of the National Cholesterol Education Program (NCEP) expert panel on detection, evaluation, and treatment of high blood cholesterol in adults (adult treatment panel III). Journal of the American Medical Association. 2001 May 16. 285(19):2486–97. doi: 10.1001/jama.285.19.2486

26. Elsayed NA, Aleppo G, Aroda VR, Bannuru RR, Brown FM, Bruemmer D, et al. Summary of Revisions: Standards of Care in Diabetes—2023. Diabetes Care. 2023 Jan 1. 46:S5–9. doi: 10.2337/DC23-SREV

27. Brugada J, Demosthenes G. Katritsis EA, Fernando Arribas JJB, Carina Blomström-Lundqvist HC, Domenico Corrado, Spyridon G. Deftereos, Gerhard-Paul Diller, Juan J. Gomez-Doblas, Bulent Gorenek, Andrew Grace, Siew Yen Ho, Juan-Carlos Kaski, Karl-Heinz Kuck PDL, Frederic Sacher GSB, et al. 2019 ESC Guidelines for the management of patients with supraventricular tachycardia The Task Force for the management of patients with supraventricular tachycardia of the European Society of Cardiology (ESC). 2020.: 655–720. doi: 10.1093/eurheartj/ehz467

28. Onder H, Yilmaz S. The Rationale of Holter Monitoring After Stroke. Angiology. 2017 Nov 1. 68(10):926–7. doi: 10.1177/0003319717703003

29. Morotti A, Goldstein JN. Diagnosis and Management of Acute Intracerebral Hemorrhage. Emergency Medicine Clinics of North America. 2016. 34(4):883–99. doi: 10.1016/j.emc.2016.06.010

30. Sharma R, Mallick D, Llinas RH, Marsh EB. Early Post-stroke Cognition: In-hospital Predictors and the Association With Functional Outcome. Frontiers in Neurology. 2020. 11(December):1–9. doi: 10.3389/fneur.2020.613607

31. Li Q, Pan S. Contrast-Associated Acute Kidney Injury: Advances and Challenges. International Journal of General Medicine. 2022. 15(February):1537–46. doi: 10.2147/IJGM.S341072

32. Smith Fawzi MC, Ngakongwa F, Liu Y, Rutayuga T, Siril H, Somba M, et al. Validating the Patient Health Questionnaire-9 (PHQ-9) for screening of depression in Tanzania. Neurology Psychiatry and Brain Research. 2019. 31(October 2018):9–14. doi: 10.1016/j.npbr.2018.11.002

33. Santangelo G, Barone P, Cuoco S, Raimo S, Pezzella D, Picillo M, et al. Apathy in untreated, de novo patients with Parkinson’s disease: validation study of Apathy Evaluation Scale. Journal of Neurology. 2014 Nov 25. 261(12):2319–28. doi: 10.1007/S00415-014-7498-1

34. Marin RS, Biedrzycki RC, Firinciogullari S. Reliability and validity of the Apathy Evaluation Scale. Psychiatry research. 1991. 38(2):143–62. doi: 10.1016/0165-1781(91)90040-V

35. Hosmer DW, Lemeshow S, Sturdivant RX. Applied Logistic Regression, 3rd Edition [Internet]. Wiley Series in Probability and Statistics. Wiley; 2013. 528 p.

36. Mellon L, Brewer L, Hall P, Horgan F, Williams D, Hickey A, et al. Cognitive impairment six months after ischaemic stroke: A profile from the ASPIRE-S study. BMC Neurology. 2015. 15(1):1–9. doi: 10.1186/s12883-015-0288-2

37. Nijsse B, Visser-Meily JMA, Van Mierlo ML, Post MWM, De Kort PLM, Van Heugten CM. Temporal evolution of poststroke cognitive impairment using the montreal cognitive assessment. Stroke. 2017. 48(1):98–104. doi: 10.1161/STROKEAHA.116.014168

38. Feigin VL, Krishnamurthi R V., Theadom AM, Abajobir AA, Mishra SR, Ahmed MB, et al. Global, regional, and national burden of neurological disorders during 1990-2015: a systematic analysis for the Global Burden of Disease Study 2015. The Lancet Neurology. 2017 Nov 1. 16(11):877–97. doi: 10.1016/S1474-4422(17)30299-5

39. Chaurasia RN, Sharma J, Pathak A, Mishra VN, Joshi D. Poststroke Cognitive Decline: A Longitudinal Study from a Tertiary Care Center. Journal of Neurosciences in Rural Practice. 2019. 10(3):459. doi: 10.1055/S-0039-1697872

40. Baccaro A, Wang YP, Brunoni AR, Candido M, Conforto AB, da Costa Leite C, et al. Does stroke laterality predict major depression and cognitive impairment after stroke? Two-year prospective evaluation in the EMMA study. Progress in neuro-psychopharmacology & biological psychiatry. 2019 Aug 30. 94. doi: 10.1016/J.PNPBP.2019.109639

41. Packheiser J, Schmitz J, Arning L, Beste C, Güntürkün O, Ocklenburg S. A large-scale estimate on the relationship between language and motor lateralization. Scientific Reports. 2020 Dec 1. 10(1). doi: 10.1038/S41598-020-70057-3

42. Hobson J. The Montreal Cognitive Assessment (MoCA). Occupational Medicine. 2015 Dec 1. 65(9):764–5. doi: 10.1093/OCCMED/KQV078

43. Banerjee G, Summers M, Chan E, Wilson D, Charidimou A, Cipolotti L, et al. Domain-specific characterisation of early cognitive impairment following spontaneous intracerebral haemorrhage. Journal of the Neurological Sciences. 2018. (2017):#pagerange#. doi: 10.1016/j.jns.2018.05.015

44. Tomlinson BE, Blessed G, Roth M. Observations on the brains of demented old people. Journal of the Neurological Sciences. 1970 Sep 1. 11(3):205–42. doi: 10.1016/0022-510X(70)90063-8

45. Kumral E, Bayam FE, Arslan H, Ph D, Orman M, Ph D. Associations Between Neuroanatomic Patterns of Cerebral Infarctions and Vascular Dementia. 2017. 1–8. doi: 10.1176/appi.neuropsych.19120356

46. Gottesman RF, Hillis AE. Predictors and assessment of cognitive dysfunction resulting from ischaemic stroke. Vol. 9, The Lancet Neurology. 2010. p. 895–905. doi: 10.1016/S1474-4422(10)70164-2

47. Caeiro L, Ferro JM, Costa J. Apathy Secondary to Stroke: A Systematic Review and Meta-Analysis. Cerebrovascular Diseases. 2013 Feb. 35(1):23–39. doi: 10.1159/000346076

48. Van Almenkerk S, Smalbrugge M, Depla MFIA, Eefsting JA, Hertogh CMPM. Apathy among institutionalised stroke patients: Prevalence and clinical correlates. American Journal of Geriatric Psychiatry. 2015 Feb 1. 23(2):180–8. doi: 10.1016/j.jagp.2014.03.011

49. Mikami K, Jorge RE, Moser DJ, Jang M, Robinson RG. Incident apathy during the first year after stroke and its effect on physical and cognitive recovery. American Journal of Geriatric Psychiatry. 2013. 21(9):848–54. doi: 10.1016/J.JAGP.2013.03.012

50. Willem Van Dalen J, Moll Van Charante EP, Nederkoorn PJ, Van Gool WA, Richard; Edo. From the Department of Neurology. 2013. doi: 10.1161/STROKEAHA

51. Rohde D, Gaynor E, Large M, Mellon L, Hall P, Brewer L, et al. The Impact of Cognitive Impairment on Poststroke Outcomes: A 5-Year Follow-Up. Journal of Geriatric Psychiatry and Neurology. 2019. 32(5):275–81. doi: 10.1177/0891988719853044

52. Lim KB, Kim J, Lee HJ, Yoo JH, You EC, Kang J. Correlation Between Montreal Cognitive Assessment and Functional Outcome in Subacute Stroke Patients With Cognitive Dysfunction. Annals of Rehabilitation Medicine. 2018 Feb 1. 42(1):26–34. doi: 10.5535/ARM.2018.42.1.26

53. Stolwyk RJ, O’Neill MH, McKay AJD, Wong DK. Are cognitive screening tools sensitive and specific enough for use after stroke?: A systematic literature review. Stroke. 2014. 45(10):3129–34. doi: 10.1161/STROKEAHA.114.004232

54. Quinn TJ, Richard E, Teuschl Y, Gattringer T, Hafdi M, O’Brien JT, et al. European Stroke Organisation and European Academy of Neurology joint guidelines on post-stroke cognitive impairment. European journal of neurology. 2021 Dec 1. 28(12):3883–920. doi: 10.1111/ENE.15068

55. Jorm AF. The Informant Questionnaire on Cognitive Decline in the Elderly (IQCODE): A review. International Psychogeriatrics. 2004 Sep. 16(3):275–93. doi: 10.1017/S1041610204000390

56. Van Rooij FG, Schaapsmeerders P, Maaijwee NAM, Van Duijnhoven DAHJ, De Leeuw FE, Kessels RPC, et al. Persistent cognitive impairment after transient ischemic attack. Stroke. 2014. 45(8):2270–4. doi: 10.1161/STROKEAHA.114.005205

57. Pendlebury ST, Wadling S, Silver LE, Mehta Z, Rothwell PM. Transient cognitive impairment in TIA and minor stroke. Stroke. 2011 Nov. 42(11):3116–21. doi: 10.1161/STROKEAHA.111.621490

58. Roebuck-Spencer TM, Glen T, Puente AE, Denney RL, Ruff RM, Hostetter G, et al. Cognitive Screening Tests Versus Comprehensive Neuropsychological Test Batteries: A National Academy of Neuropsychology Education Paper. Archives of Clinical Neuropsychology. 2017 Jun 1. 32(4):491–8. doi: 10.1093/ARCLIN/ACX021

